# Diabetes, impaired fasting glucose, and cognitive trajectories: a multi-cohort study

**DOI:** 10.64898/2026.05.26.26354185

**Authors:** Jessica W. Lo, John D. Crawford, Katherine Samaras, Richard B. Lipton, Mindy J. Katz, Carol A. Derby, Pierre-Marie Preux, Maëlenn Guerchet, Eleonora d’Orsi, Anna Quialheiro, Cassiano Ricardo Rech, Karen Ritchie, Elena Rolandi, Annalisa Davin, Michele Rossi, Suzana Shahar, Norfadilah Rajab, Nurul Fatin Malek Rivan, Mary Ganguli, Erin Jacobsen, Beth E. Snitz, Henry Brodaty, Yen-Ching Chen, Jen-Hau Chen, Matthew Lennon, Darren M. Lipnicki, Perminder S. Sachdev

## Abstract

**INTRODUCTION:** Cognitive trajectories may clarify how type 2 diabetes (T2D) and impaired fasting glucose (IFG) relate to dementia risk, but longitudinal associations remain unclear, particularly in the context of stroke.

**METHODS:** Data from 5,631 dementia- and stroke-free older adults (mean age 75 years) from 7 international population-based cohorts were analyzed. Linear mixed-effects models estimated cognitive trajectories during stroke-free and post-stroke follow-up. Glucose status was defined by fasting glucose and prior T2D diagnosis.

**RESULTS:** Over 6.6 years of follow-up (4.5% with incident stroke), T2D was associated with lower baseline cognitive performance compared with normal fasting glucose (−0.14 SD, 95% CI −0.21 to −0.07), but not with faster cognitive decline during stroke-free or post-stroke follow-up. IFG was not associated with lower cognitive performance or faster decline.

**DISCUSSION:** In older adults, T2D was associated with persistently lower cognitive performance but not faster decline, suggesting adverse cognitive effects may be established before late life.

## 1 BACKGROUND

Type 2 diabetes (T2D) affects approximately one in five adults aged 65–99 years globally, and its prevalence is increasing with population ageing [1]. Beyond its established role in cardiovascular disease, including stroke, and mortality [2], T2D is recognized as a major risk factor for dementia [3,4]. Diabetes may contribute to cognitive impairment through vascular and metabolic pathways that lead to structural brain changes [5].

However, associations between T2D and cognitive decline are generally more modest and variable than those reported for dementia, with findings differing across cognitive domains, age groups, and study designs [5–8]. Evidence for prediabetes or impaired fasting glucose (IFG) is also limited and mixed, with some studies finding no association and others reporting modest effects [9–12]. Prior studies have mostly focused on dementia as a clinical endpoint, which may obscure earlier or more subtle cognitive differences. Examining cognitive trajectories, including both overall level of cognitive performance and rate of cognitive decline over follow-up, may clarify whether cognitive differences associated with T2D and IFG are already present by older age, whether they continue to widen over follow-up, or differ across cognitive domains.

Associations between glucose status and cognitive trajectories may also be influenced by stroke, which is common in older adults and a major determinant of cognitive decline and dementia [13,14]. Because T2D increases stroke risk and stroke survivors with T2D have poorer cognitive outcomes [15], it is important to account for incident stroke over follow-up and examine whether T2D or IFG are associated with additional cognitive decline beyond that attributable to stroke itself.

In this pooled analysis of harmonized data from population-based cohorts in the Cohort Studies of Memory in an International Consortium (COSMIC) [16], we examined whether older adults with T2D, IFG, and normal fasting glucose differed in cognitive performance and rates of cognitive decline, and whether these associations differed before and after incident stroke.

## 2 METHODS

### 2.1 Sample and baseline characteristics

Nine COSMIC cohorts with fasting glucose level (FGL) and incident stroke data over follow-up were included (Table 1). Participants with a history of stroke or dementia at baseline were excluded (eTable 1 in the Supplement). Stroke was self-reported in all studies except one, in which it was identified by clinical examination (eTable 2).

**Table 1.**
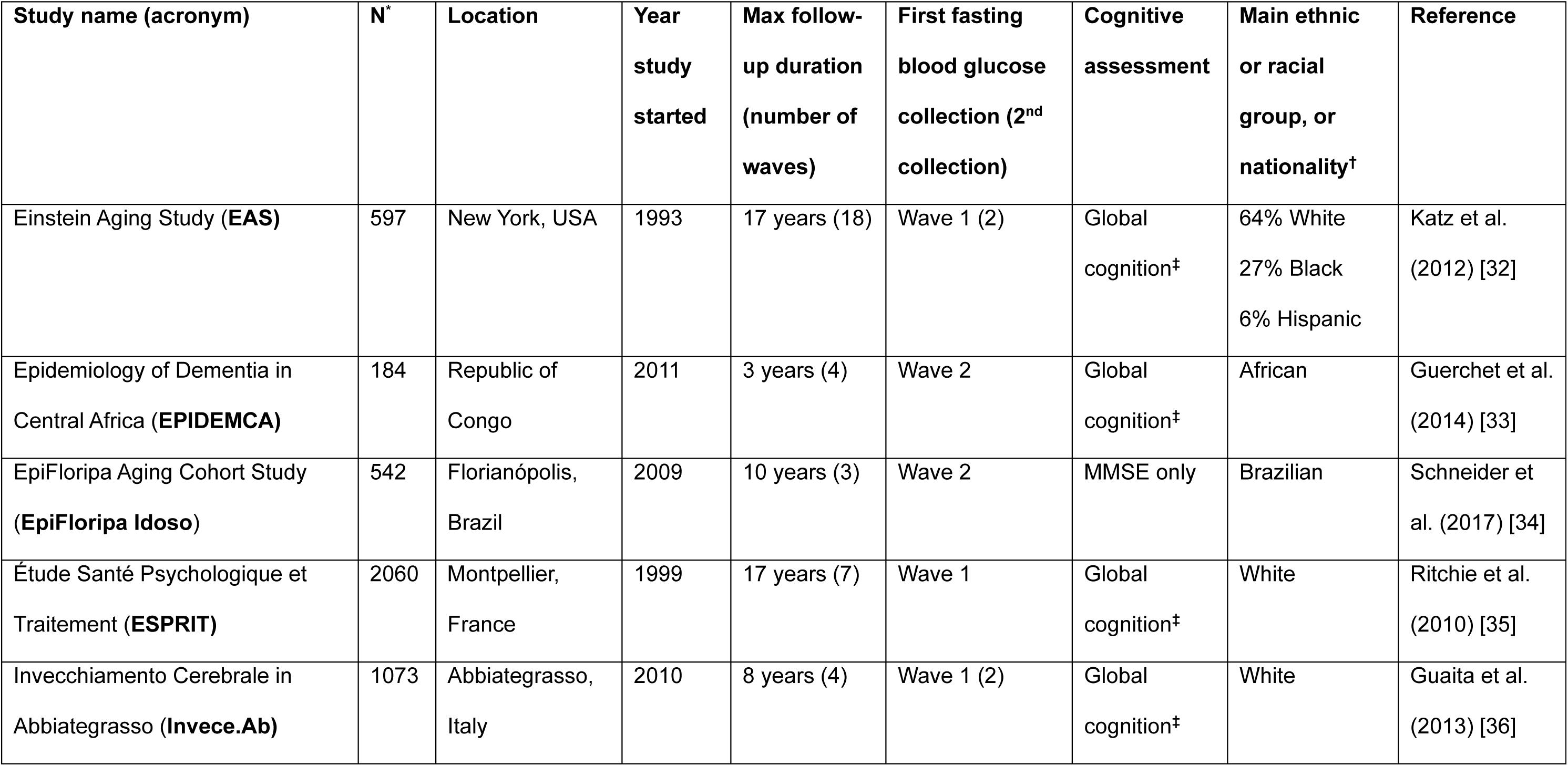
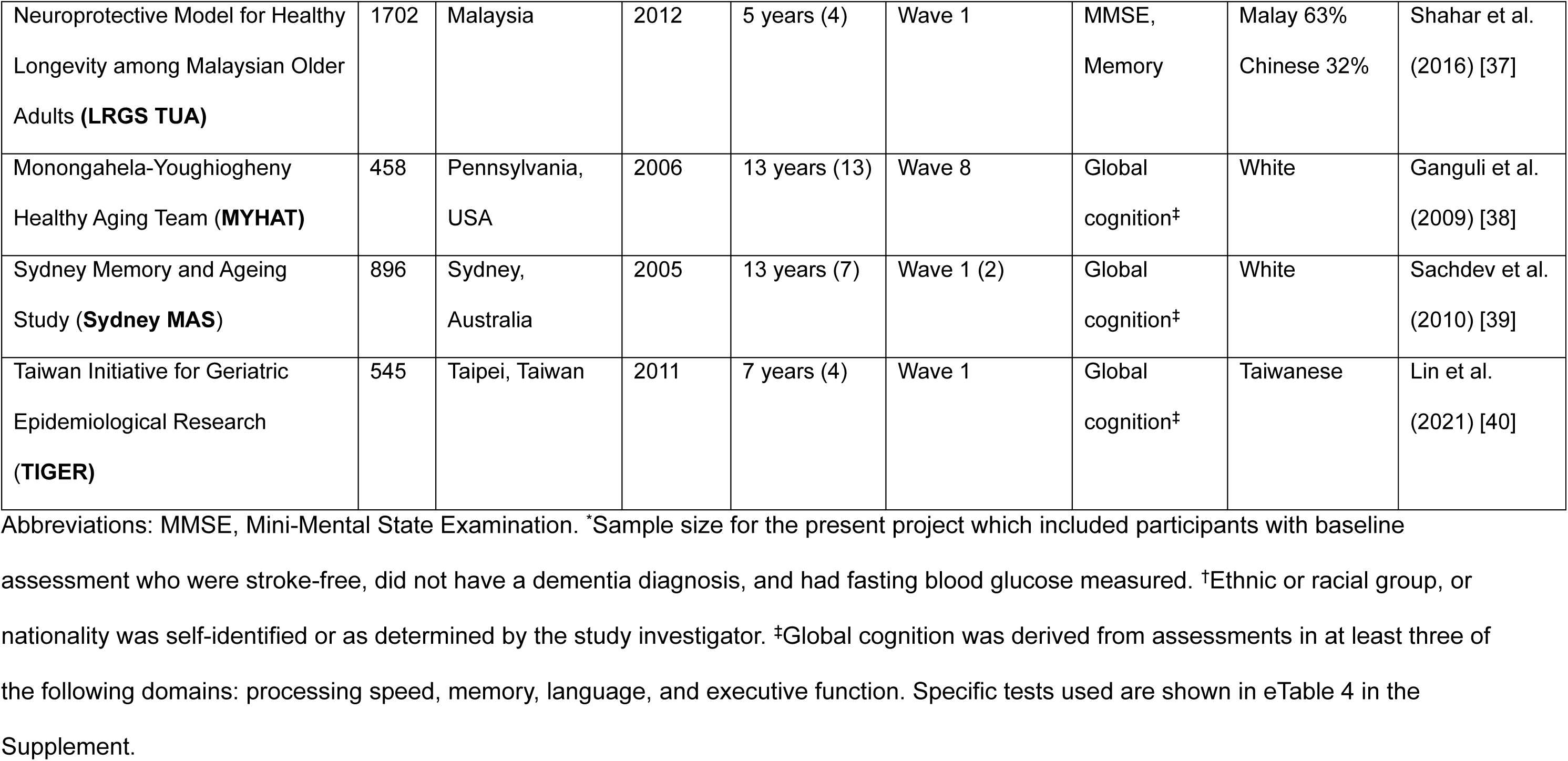
Contributing study cohorts.

Baseline (wave 1) covariates were harmonized according to previous COSMIC protocols [13,17] and included age, sex, years of education, race, apolipoprotein E ε4 carrier status (APOE4), blood pressure, body mass index (BMI), smoking (ever), alcohol use (>1 vs ≤1 drink/week), physical activity (minimal vs moderate-to-vigorous), depression, hypertension, hypercholesterolemia, and cardiovascular disease (CVD). To account for between-cohort differences in educational systems and average years of education, a study-level mean education variable was included. Race was included because of known disparities in diabetes and cognitive outcomes and to account for between-cohort heterogeneity in ethno-racial composition. Because of small subgroup sizes, racial categories were collapsed into White and a combined category comprising Asian, African, Brazilian, Hispanic, and other groups. Harmonization procedures are described in eMethods 1 and eTable 3 in the Supplement.

### 2.2 Glucose status

Fasting glucose level (FGL) measurements were collected at the baseline study visit (wave 1) in six cohorts and at later waves in three cohorts (Table 1). Glucose status was classified using American Diabetes Association criteria [18]. Specifically, type 2 diabetes (T2D) was defined as FGL ≥7.0 mmol/L or prior diabetes diagnosis or treatment at or before study entry. Impaired fasting glucose (IFG) was defined as FGL 5.6–6.9 mmol/L, and normal fasting glucose (NFG) as FGL <5.6 mmol/L, both in the absence of prior diabetes diagnosis or treatment. Prior diabetes diagnosis was determined from medical history or self-report (eTable 3).

Some individuals with a prior diabetes diagnosis may have had type 1 diabetes; however, differentiation was not possible. Type 1 diabetes accounts for approximately 5–10% of all diabetes cases overall and a smaller proportion among older adults [19]; therefore, any resulting misclassification is likely to have minimal impact.

### 2.3 Cognitive outcome

Neuropsychological assessments were conducted longitudinally. For each cohort and cognitive domain (processing speed, memory, language, and executive function), the most commonly administered test, or a close equivalent, was selected (eTable 4). The visuospatial domain was not included because there was insufficient overlap in visuospatial tests across cohorts.

Scores were standardized using a demographically centered approach [20], based on predicted baseline cognitive performance for the average participant in the combined sample, consistent with previous COSMIC work [13,17] (eTable 5; eMethods 2). Global cognition, defined as the standardized mean of at least three domains, was available in seven cohorts. In cohorts where FGL was first collected after study baseline, only cognitive assessments obtained after the first FGL collection were included in the analyses.

### 2.4 Statistical Methods

#### 2.4.1 Descriptive statistics

Baseline characteristics were compared across glucose groups and by follow-up status using *t* tests or χ² tests, with effect sizes quantified using Cohen’s *d* or *h,* for continuous and categorical data, respectively.

#### 2.4.2 Primary analysis with global cognition

One-step Individual Participant Data (IPD) meta-analysis was used to analyze data from all studies simultaneously within a hierarchical mixed-effects framework. A two-step approach was not used because one-step IPD models are well suited to modelling longitudinal trajectories and are preferred when interaction effects are of primary interest [21,22]. Global cognition was used as the primary outcome.

To account for potential effects of incident stroke during follow-up on cognitive trajectories, analyses used a regression discontinuity framework with two sequential linear mixed-effects models, with incident stroke as the discontinuity event [23], consistent with our previous COSMIC analysis [13]. Models included time in study (TIS; rate of cognitive change during stroke-free follow-up), a time-varying stroke indicator (acute cognitive change at stroke onset), and time since stroke (TSS; change in post-stroke slope relative to TIS), together with glucose status and glucose-by-time interaction terms.

The basic model included these terms together with baseline age, sex, and education. Interactions with age and sex were tested and retained if p < 0.10. Additional interactions between glucose status and stroke-related terms were tested to assess whether associations differed before or after incident stroke; interaction terms (p < 0.10) were retained in the model.

For the fully adjusted model, potential baseline covariates were first added individually to the basic model, and those associated with global cognition at p < 0.10 were included as potential confounders. The fully adjusted model therefore included all terms from the basic model together with race, study-level mean education, and selected potential confounders (see Results 3.1 and eTable 11).

Model intercepts represent adjusted cognitive performance at the start of follow-up, whereas TIS represents the rate of cognitive decline in the NFG group during stroke-free follow-up; interaction terms with these variables represent differences in cognitive performance and decline for IFG and T2D relative to NFG.

In all models, random intercepts for study and participant were included to account for clustering of repeated measures within individuals and clustering of participants within cohorts. Models were estimated using restricted maximum likelihood. Missing covariates were imputed using multiple imputation by chained equations (eMethods 3; eTable 6). Predicted trajectories were plotted at mean covariate values (eTable 7).

#### 2.4.3 Sensitivity analyses and secondary analyses

Two sensitivity analyses were conducted for the primary analysis. First, analyses were restricted to participants with complete data. Second, we excluded the cohorts in which fasting glucose measurements were not collected at study baseline, which required exclusion of earlier cognitive assessments from the primary analysis.

Secondary analyses repeated the primary model using domain scores and MMSE as outcomes. Two additional analyses were conducted in subgroups with follow-up FGL measurements (1–2 years after the initial measurement) and global cognition data.

First, glucose transition groups (stable normal, stable IFG, IFG to normal, IFG to T2D, normal to IFG or T2D, and baseline T2D regardless of transition) were used in place of baseline glucose status to examine associations between changes in glucose status and cognitive decline. Second, average FGL was calculated from the two measurements, and glucose status was redefined based on the average FGL.

Finally, secondary exploratory analyses examined metformin use in subgroups with available data. The T2D group was subdivided into participants with and without reported metformin use to explore whether metformin use influenced findings within the T2D group. Exploratory analyses focused on metformin because harmonized data on other glucose-lowering medications were limited across cohorts.

All analyses were conducted using Stata version 18.0 (StataCorp).

### 2.5 Ethics approval and reporting

This study was approved by the University of New South Wales Human Research Ethics Committee (HC12446 and HC17292). Each cohort received local ethics approval (eTable 8), and all participants provided informed consent. Reporting followed the Strengthening the Reporting of Observational Studies in Epidemiology (STROBE) guidelines [24].

## 3 RESULTS

Among 8057 participants from nine cohorts, 4704 (58%) had normal fasting glucose, 1636 (20%) had IFG, and 1717 (21%) had T2D. Half were followed to study completion or had more than 11 years of follow-up; participants lost to follow-up were older and had a higher burden of vascular risk factors, although most standardized differences were small (eTable 9). Compared with participants with normal fasting glucose, those with IFG or T2D were more often male, had lower educational attainment, and had a higher prevalence of vascular risk factors, although most standardized effect sizes were small (Table 2).

**Table 2.** Baseline characteristics of glucose status groups.

|  | **Type 2**<br>**Diabetes**<br>**(T2D)** | **Impaired**<br>**Fasting**<br>**Glucose**<br>**(IFG)** | **Normal**<br>**Fasting**<br>**Glucose**<br>**(NFG)** | **P-value\***<br>**(Cohen *d* or**<br>**Cohen *h*)**<br>**(T2D vs NFG)<sup>†</sup>** | **P-value\***<br>**(Cohen *d* or**<br>**Cohen *h*)**<br>**(IFG vs NFG)<sup>†</sup>** |
| --- | --- | --- | --- | --- | --- |
| N, % | 1717 (21) | 1636 (20) | 4704 (58) | - | - |
| Age, y | 71.8 (6.4) | 73.0 (6.1) | 72.8 (6.2) | <0.001 (0.17) | 0.38 (0.03) |
| Male sex, % | 855 (50) | 821 (50) | 1783 (38) | <0.001 (0.24) | <0.001 (0.24) |
| Education, y | 8.4 (5.0) | 9.0 (5.0) | 9.8 (4.9) | <0.001 (0.29) | <0.001 (0.17) |
| White participants<sup>‡</sup> | 758 (44) | 1056 (65) | 3035 (65) | <0.001 (0.41) | 0.98 (0.002) |
| Body mass index | 27.6 (5.0) | 27.0 (4.7) | 25.2 (4.2) | <0.001 (0.53) | <0.001 (0.40) |
| APOE4 carrier | 175 (19) | 239 (21) | 685 (20) | 0.50 (0.03) | 0.80 (0.02) |
| Systolic blood pressure | 140.7 (20) | 141.2 (20) | 138.5 (19) | <0.001 (0.12) | <0.001 (0.14) |
| Diastolic blood pressure | 78.3 (12) | 80.2 (11) | 78.6 (11) | 0.37 (0.03) | <0.001 (0.15) |
| Hypertension | 1421 (83) | 1196 (73) | 3129 (67) | <0.001 (0.37) | <0.001 (0.13) |
| High cholesterol | 806 (48) | 665 (41) | 1973 (42) | <0.001 (0.12) | 0.37 (0.02) |
| Cardiovascular disease | 426 (26) | 363 (23) | 843 (18) | <0.001 (0.19) | 0.010 (0.12) |
| Smoker (ever) | 705 (41) | 712 (44) | 1742 (37) | 0.003 (0.08) | <0.001 (0.14) |
| Alcohol use (at least<br>one drink p/w) | 515 (47) | 804 (60) | 2217 (59) | <0.001 (0.24) | 0.34 (0.02) |
| Physical activity<br>(moderate/vigorous) | 762 (69) | 981 (75) | 2892 (78) | <0.001 (0.20) | 0.008 (0.07) |
| History of depression | 301 (18) | 304 (19) | 1164 (25) | <0.001 (0.17) | <0.001 (0.15) |

Data are presented as mean (SD) or n (%). \*Pearson's $\chi^2$ tests and t tests were used to
examine group differences. <sup>†</sup>For both Cohen's *d* and *h*, values of 0.2, 0.5, and 0.8 are
considered to represent small, medium, and large differences, respectively, between groups
in means or proportions. <sup>‡</sup>Because of small subgroup sizes, racial categories were collapsed
into White and a combined category comprising Asian, African, Brazilian, Hispanic, and other
groups.

### 3.1 Primary analyses with global cognition

The primary analysis included 5631 participants from seven cohorts with available global cognition data (mean [SD] age, 74.8 [5.8] years; 58% female; mean follow-up, 6.6 [4.6] years) (eFigure 1). Within this sample, 251 participants (4.5%) experienced an incident stroke (55% with NFG, 21% with IFG, and 24% with T2D), occurring a mean (SD) of 4.5 (3.5) years after baseline.

**Figure 1.**
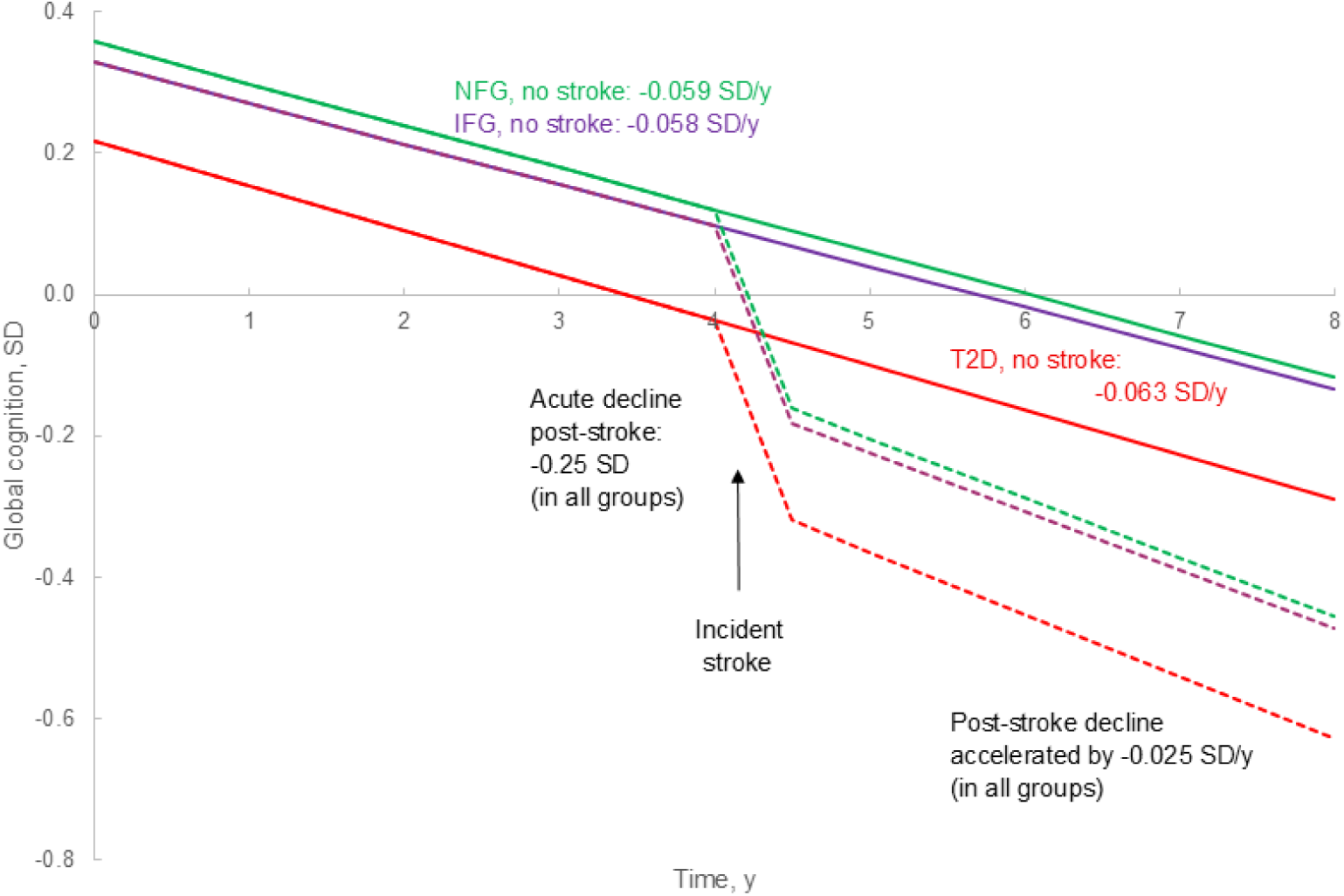
Projected values of global cognition without/before and after stroke by glucose status group (note, color should be used in print) Red line = T2D, type 2 diabetes; purple line = IFG, impaired fasting glucose; green line = NFG, normal fasting glucose. Solid lines denote trajectories during stroke-free follow-up; dotted lines denote post-stroke trajectories. N = 5,631 participants with global cognition. Predicted global cognition values were calculated using common baseline covariate values (eTable 7). Incident stroke was modeled as occurring 4.5 years after study baseline, corresponding to the mean time to incident stroke in the sample.

In the final adjusted model, global cognition at baseline was lower in T2D (−0.14 SD; 95% CI −0.21 to −0.069) but did not differ significantly for IFG (−0.029 SD; 95% CI −0.094 to 0.036) compared with normal fasting glucose (Table 3). Neither IFG nor T2D was associated with faster cognitive decline compared with NFG (Table 3).

**Table 3.** Adjusted estimates of differences in baseline level and rate of decline in global cognition over time by glucose status group.

| Measure (model variable) | Coefficient (95% CI) | P-value |
| --- | --- | --- |
| *Stroke-free follow-up* |  |  |
| Baseline level / intercept (glucose status) |  |  |
| IFG | -0.029 (-0.094, 0.036) | 0.38 |
| T2D | -0.14 (-0.21, -0.069) | **<0.001** |
| Slope (TIS; SD/y) | -0.059 (-0.062, -0.056) | **<0.001** |
| Moderating effect of glucose status on slope<br>(TIS × glucose status) |  |  |
| TIS × IFG | 0.0016 (-0.0049, 0.0082) | 0.63 |
| TIS × T2D | -0.0041 (-0.012, 0.0032) | 0.27 |
| *Post-stroke follow-up* |  |  |
| Acute effect of stroke on cognitive level<br>(stroke; SD) | -0.25 (-0.35, -0.15) | **<0.001** |
| Difference in post-stroke slope relative to<br>before stroke (TSS; SD/y) | -0.025 (-0.047, -0.0020) | **0.033** |

Abbreviations: TIS, time in study; TSS, time since stroke; T2D, type 2 diabetes; IFG, impaired fasting glucose.
N = 5,631 participants with global cognition data from 7 studies. Estimates are from the final fully adjusted model, which included age, sex, education, race, high cholesterol, APOE ε4 status, smoking, alcohol use, physical activity, history of depression, and the age × TIS and sex × TIS interactions. The reference group was normal fasting glucose. Bold values indicate $p < 0.05$ .

Estimates were similar in the fully adjusted and basic models, with effect estimates unchanged or differing by less than 13% (eTable 10). The age × TIS and sex × TIS interactions were significant and retained. The fully adjusted model additionally included high cholesterol, alcohol use, physical activity, smoking, depression, and APOE4 (eTable 11).

Consistent with our previous COSMIC analysis [13], incident stroke was associated with both an acute decline in cognition and accelerated subsequent decline (Table 3). However, interactions between glucose status and stroke-related cognitive change were not significant and were therefore not retained in the model (eTable 12). Rates of cognitive decline during stroke-free follow-up were also similar between participants who later experienced incident stroke and those who remained stroke-free, with no statistically significant differences (eTable 13).

Figure 1 illustrates estimated cognitive trajectories for the glucose groups over stroke-free and post-stroke follow-up. Cognitive performance in T2D remained consistently lower across follow-up, whereas cognitive trajectories were very similar for IFG and NFG.

### 3.2 Sensitivity and secondary analyses

Sensitivity analyses using complete-case data and excluding the two cohorts in which fasting glucose was not measured at baseline yielded consistent results; the direction and interpretation of effects were unchanged (eTable 14).

Domain-specific analyses showed similar patterns across all four domains: T2D was associated with persistently lower cognitive performance across follow-up by 0.10–0.14 SD, but not faster cognitive decline, compared with NFG (eTable 15; eFigure 2). Cognitive performance and rates of decline for IFG did not differ significantly from those for NFG across all domains.

Although MMSE slopes differed between groups, baseline MMSE performance did not, and slope differences were small (≤ 0.054 points/year; eTable 15; eFigure 2) and well below thresholds considered clinically meaningful (2–4 points over 1.5 years) [25]. Overall MMSE decline was 0.046 points/year (95% CI −0.056 to −0.036), suggesting limited sensitivity to change.

Four cohorts (n = 2134) provided follow-up fasting glucose measurements (eTable 16A). Glucose status remained stable in 53% of participants; 10% reverted from IFG to normal glucose, and 9% progressed from normal glucose to IFG or T2D (eTable 16B). Findings were broadly consistent with the primary analysis, with lower cognitive performance in T2D and near-zero differences in cognitive decline between groups (eTable 16C). Results were also similar when glucose groups were redefined using the average of two fasting glucose assessments (eTable 17).

Four cohorts also collected data on metformin use, with 2441 participants included in this analysis (eTable 18A). Among participants with T2D (n = 510, 21%), 242 (47%) reported metformin use. Patterns were similar to those in the primary analysis: both T2D groups, with and without metformin use, had lower cognitive performance across follow-up, whereas rates of cognitive decline were similar across groups (eTable 18B).

## 4 DISCUSSION

In this pooled analysis of nine population-based cohorts of older adults, T2D was associated with persistently lower cognitive performance throughout follow-up. Neither T2D nor IFG was associated with faster cognitive decline, and IFG showed cognitive performance and trajectories similar to those of normal fasting glucose. These findings were consistent across global cognition, processing speed, memory, language, and executive function and were robust to adjustment for vascular and other risk factors associated with cognitive decline. There was no evidence that incident stroke altered the associations between glucose status and cognitive trajectories.

### 4.1 T2D and cognitive trajectories

Previous studies consistently report that T2D is associated with cognitive dysfunction and increased dementia risk [5,9,26]. However, evidence for accelerated cognitive decline has been more variable across cognitive domains and age groups [5,8,27]. For example, a Swedish cohort with a mean age of 63 years reported domain-specific accelerated decline [6], whereas a Dutch study of adults aged 85 years or older found lower baseline cognitive functioning in individuals with diabetes but no accelerated cognitive decline [7]. Our findings are more consistent with studies of older adults, as T2D was associated with persistently lower cognitive performance but not faster decline.

This pattern may indicate that cognitive differences associated with T2D were already present by older age, rather than continuing to widen during late-life follow-up. The cognitive effects of T2D may develop gradually over decades, such that lower cognitive performance observed in later life could reflect vascular and metabolic injury that accumulated before study entry. However, we cannot determine whether lower cognitive performance in T2D reflected accumulated effects of diabetes, pre-existing differences before diabetes onset, or both. Future studies spanning mid- to late life are needed to determine how cognitive trajectories associated with T2D evolve across different stages of ageing, and how these associations vary according to disease duration and age at diabetes onset.

Survivor bias may also have contributed, as individuals with more advanced diabetes-related morbidity or cognitive impairment may have been less likely to participate, remain in follow-up, or may have died from diabetes-related vascular disease before study participation or follow-up. Lower fasting glucose in late life may also reflect frailty or comorbidity in some individuals, which could contribute to cognitive decline in the NFG reference group and weaken differences in cognitive trajectories between T2D and NFG. Heterogeneity within the T2D group, including variation in disease duration, glycemic control, and treatment, may also have weakened associations with cognitive decline. We found no evidence that cognitive trajectories differed between T2D subgroups with and without metformin use; however, these analyses may have been underpowered, and we could not account for other glucose-lowering medications, glycemic control, or diabetes duration because these data were unavailable across most cohorts.

### 4.2 IFG and cognitive trajectories

Fewer studies have examined the association of IFG with cognitive decline and dementia, and findings have been inconsistent. Some studies using cumulative or average glucose exposure have reported increased dementia risk or faster decline with higher glucose or HbA1c levels in individuals without diabetes or with normoglycemia [9,10,28]. However, these studies either included younger participants, with mean ages around 50 or 66 years, or examined dementia outcomes rather than cognitive decline.

In contrast, studies based on single fasting glucose measurements in older adults have generally found no association with dementia risk [4] or cognitive decline [11,12,27], although one study reported lower baseline memory but slower subsequent decline in individuals with IFG [6].

In our study, IFG was not associated with lower cognitive performance or faster decline than normal fasting glucose. Although IFG was defined using a single fasting glucose measurement and therefore may not reflect long-term glycemic exposure, findings were directionally similar in the subset with follow-up fasting glucose measurements. However, the smaller sample size in this subgroup may have limited power to detect differences.

Although cumulative glycemic exposure has been linked to higher dementia risk or faster cognitive decline in midlife and early older age, it remains unclear whether this association persists in older adults. Some studies have reported that progression from prediabetes to diabetes is associated with accelerated cognitive decline [29,30], although this was not observed in our smaller subset and warrants further investigation. Future studies should clarify whether cumulative glycemic exposure contributes to cognitive decline in older adults.

The similarity in trajectories between IFG and normal fasting glucose may also suggest that, in older adults, remaining free of T2D into later life reflects a more favorable cumulative metabolic profile. Lifestyle or treatment changes following identification of elevated glucose may also have contributed to a return to normal fasting glucose over follow-up in some individuals.

### 4.3 Cognitive domains and post-stroke cognitive trajectories

Across cognitive domains, findings were similar to those for global cognition. T2D was associated with lower performance but not faster decline across all domains, whereas IFG was similar to normal fasting glucose in both level and rate of decline. Although some studies have more consistently implicated memory, processing speed, and executive function than language in diabetes-related cognitive dysfunction [5,7,27], our findings suggest that, in older adults, the cognitive effects of T2D may be broad rather than confined to specific domains.

Previous studies have reported poorer post-stroke cognitive outcomes in diabetes but not prediabetes [15,31]. To our knowledge, this is the first study to examine IFG and T2D in relation to cognitive trajectories before and after incident stroke.

We found no evidence that T2D or IFG was associated with accelerated cognitive decline before or after stroke. However, the smaller numbers of participants with incident stroke within each glucose group may have limited power to detect differences. Attrition may also have attenuated differences in post-stroke cognitive trajectories between glucose groups, as participants with T2D or IFG who experienced severe stroke or poorer outcomes may have been less likely to return for follow-up assessments. Even without evidence of additional diabetes-related acceleration after stroke, lower cognitive performance before stroke in T2D, together with the acute and long-term cognitive decline associated with stroke itself, may mean that stroke survivors with T2D reach clinically significant cognitive impairment or dementia earlier.

### 4.4 Strengths, limitations and conclusion

Strengths of this study include the large sample of more than 5,600 individuals from seven international population-based cohorts in the primary analysis, which increased statistical power to examine cognitive trajectories and interactions across glucose groups, the relatively long follow-up, adjustment for key vascular risk factors, examination of multiple cognitive domains, and the use of longitudinal models that accounted for stroke-related cognitive change when evaluating cognitive trajectories. The inclusion of diverse international populations also supports the generalizability of the findings.

This study also has several limitations. The number of participants with incident stroke in each glucose group was small, limiting power to detect differences in stroke-related cognitive change. Glucose status was based on a single fasting glucose measurement at baseline, which may not reflect long-term glycemic exposure. HbA1c and other measures of cumulative dysglycemia were unavailable for most cohorts and may better capture chronic glycemic burden. Data on diabetes duration and glucose-lowering medication use were not available across most studies, limiting our ability to examine whether disease chronicity or treatment influenced cognitive trajectories. Attrition was substantial, as is common in studies of older adults, and individuals with poorer health or cognition may have been less likely to complete follow-up, potentially biasing findings toward the null. Harmonization across international cohorts meant that only the most commonly administered test was used to represent each cognitive domain, which may have reduced precision and sensitivity to subtle domain-specific differences, and residual between-cohort heterogeneity may have increased variability in the estimates.

In this pooled analysis of international population-based cohorts, T2D was associated with persistently lower cognitive performance but not faster cognitive decline, whereas IFG was associated with cognitive trajectories similar to those of normal fasting glucose. These findings suggest that the adverse cognitive effects of T2D may be established before late life and highlight the importance of strategies across adulthood to prevent diabetes and preserve metabolic and brain health.

## Data Availability

All data produced in the present study may be accessed upon an application via the COSMIC consortium.

## Abbreviations

T2D: type 2 diabetes
IFG: impaired fasting glucose
FGL: fasting glucose level
NFG: normal fasting glucose
IPD: individual participant data
TIS: time in study
TSS: time since stroke.

## ACKNOWLEDGEMENTS

We thank the participants and their informants for their time and generosity in contributing to this research. We also thank the research teams from each contributing study. The TIGER study is grateful for the technical support provided by the Sequencing and Biochemistry Core, Department of Medical Research, National Taiwan University Hospital. The InveCe.Ab team is grateful to Antonio Guaita, the former PI of the study.

The research scientific committee of COSMIC, comprised of principal investigators of study cohorts, leads the scientific agenda of the consortium and we thank them for ongoing support and governance. The list of COSMIC research scientific committee and additional principal investigators can be found at https://cheba.unsw.edu.au/consortia/cosmic/scientific-committee.

JW Lo acknowledges the use of ChatGPT 5.5 (accessed April–May 2026) to assist with reorganization of the Introduction and Discussion sections for clarity and flow, and to help revise the manuscript in response to co-author comments. All AI-generated suggestions were reviewed and revised by JW Lo, and the final manuscript was reviewed and approved by all authors.

