## Supplemental files for "Diabetes, impaired fasting glucose, and cognitive trajectories: a multi-cohort study"

### SUPPLEMENTARY METHODS

#### eMethods 1. Data categorization

Sex was coded as Male or Female.

Ethnic, racial, or nationality group was self-identified or determined by the principal investigators as the predominant group in the cohort. They were initially classified into categories including White, Asian, Black American, African, Brazilian, Mexican, Hispanic American, and Other. Because of small subgroup sizes, categories were collapsed into White and all other racial groups for the analysis.

Alcohol use was initially categorized as 0–never drinks or <1 drink per week, 1–minimal or 1 drink per week; 2–2 or more drinks per week. The key model included the binary categorization: 0–never drinks or <1 drink per week, 1–more than 1 drink per week.*

Physical activity was initially coded as 0–minimally active; 1–moderate activity at least once a week; 2–vigorous activity at least once a week. The key model included the binary categorization 0–minimally active, 1–moderate to vigorous activity.*

Smoking was coded as 0–never smoker; 1–ever smoker (current or past), since 5 studies coded smoking in this way and did not have data on current smoking status.

Study entry period was coded as 0–before 1990, 1–1990s, 2–2000 onwards.

History of diabetes, hypertension, high cholesterol, and cardiovascular disease were categorized as yes/no. Methods of harmonization are presented in eTable 3.

Apolipoprotein E ε4 allele (APOE4) carrier was coded as 0–not a carrier, 1–at least 1 APOE4 allele.

Depression was defined as current depressive symptoms or depression at baseline. See eTable 3 for definition used in each study.

Continuous variables were used as provided by each study: age at baseline, education in years, measurement of systolic and diastolic blood pressure at baseline, BMI (calculated as weight in kg/[height in m]$^2$).

*The categories in each of these factors were separately collapsed into two to allow convergence in the multiple imputation of the covariates.

#### eMethods 2. Standardization of neuropsychological tests and computation of domain z-scores

One approach to harmonizing neuropsychological test scores is via the calculation of standardized scores. As described by Griffith et al 2016, there are two commonly used standardization methods. The first and more common method involves the calculation of standardized scores as Z-scores (or any linear transformation of these, such as T-scores) using adjusted means and standard deviations (SDs) based on a reference sample. The adjusted means are the residuals in a regression model, with test score as the dependent variable and age, sex, and education as the predictors. This approach is commonly used when test scores need to be evaluated relative to demographically similar individuals so that impairment status can be assessed. However, it is difficult to examine the effects of sex, age and education on cognitive function using this approach since scores are adjusted for these factors. Therefore, another approach to harmonizing test scores from multiple cohorts using standardization has been developed, which Griffith et al called ‘category-centered scores’ [Griffith et al 2016].

This approach is an extension of the use of the mean and SD from a reference sample to calculate standardized scores for individuals in the patient group or some other sample. However, when analyzing data from multiple cohorts and different reference groups, then the standardized scores would not be comparable across the different studies since each reference group would have different distributions of age, sex, and education. The solution, as described in Griffith et al, is to select subsamples from within each reference group with the same overlapping ranges of demographic characteristics and use the means and SDs of these subsamples to obtain standardized scores.

A variation of this procedure is to obtain separate regression estimates of the means and SDs at the same common values of demographics from each of the reference groups. This minimizes the risk that the ranges of demographic characteristics in the subsamples might be much smaller in size than the original samples, which would cause the estimation of means and SDs to be less reliable.

In longitudinal research, when the baseline control/reference sample is deemed to be a representative sample of the relevant population, the baseline sample is commonly used as the reference group to obtain standardized scores for both baseline and later assessment waves.

Using the regression-based approach for the calculation of category-centered scores, the procedure is as follows:

1. Calculate the mean or median values of age, sex, and education in the combined sample from all cohorts. Make sure that these mean (or median) values lie well within the range (and not outside or at the edges) of each cohort. If not, shift the values.

2. In each cohort, build a regression model with the raw test score at baseline as the dependent variable, and age, sex, and education as predictors. Using the ‘common values’ of age, sex and education, as determined in step 1, calculate the estimated mean test score. The estimated SDs are the unbiased estimates of the SDs of the residuals or the standard deviation of the residual in this model and can be obtained as the Standard Errors of the Estimates in the output of the regression analysis.

3. Calculate the standardized test scores for all waves in each cohort using the estimated mean and SD as calculated in step 2.

##### Step-by-step protocol

1. We chose the common values of age, sex and education as: 73 (age), 0.43 (sex, where 1 is male, 0 is female), 10.0 (education in years).
2. For a particular study cohort, we constructed regression model with raw test scores at baseline as the dependent variable, and age, sex, and education as predictors.
3. We calculated estimated mean test score for the ‘common’ individual based on the model estimates and common values as described above in step 1.

4. Based on the estimated SD of the residuals (in STATA output, it is the root MSE) from the regression model from step 2 and the estimated test score estimated in step 3, we calculated standardized test scores for all waves.
5. Based on the assignment of test to a domain, we renamed the test as the domain.
6. Global cognition was calculated as the mean of four, or at least three, cognitive domains and then divided by its SD.

#### eMethods 3. Multiple imputation

We assumed that missing data were missing at random (MAR) and multiple imputation (MI) using chained equations was used to impute missing covariates. All baseline covariates including baseline global cognition and domain scores, as well as demographic data were included in the imputation model. Follow-up data were not included in the imputation because each study and participant have different follow-up schedules. Since some covariates (APOE4, CVD, high cholesterol, and physical activity) were missing completely in some studies, imputation was performed in the full sample. Since the clustering of studies was not the focus of our analysis, we ignored clustering of studies in the imputation model for simplicity and only allowed for it in the analysis model [White et al. 2011].

The fraction of incomplete data in the adjusted model was about 20 per cent, hence we used 20 imputations as recommended in [White et al. 2011].

### SUPPLEMENTARY TABLES

##### eTable 1. Dementia diagnosis criteria at study entry and information about each included study cohort

| Study name | Study acronym | Diagnostic criteria for dementia at baseline * | About study/ Recruitment strategies ** |
|---|---|---|---|
| Einstein Aging Study | **EAS** | DSM-IV | Between 1993 and 2004, Health Care Financing Administration/ Centers for Medicaid and Medicare Services (HCFA/CMS) rosters of Medicare eligible persons aged 70 and above were used to develop sampling frames of community residing participants in Bronx County in the US. Since 2004, New York City Board of Elections registered voter lists for the Bronx have been used due to changes in policies for release of HCFA/CMS rosters. |
| Epidemiology of Dementia in Central Africa | **EPIDEMCA** | DSM-IV and NINCDS-ADRDA criteria | A multicenter population-based study was carried out in Central African Republic and Republic of Congo between 2011 and 2012 including both urban and rural sites in each country. Follow-up was performed in Congo only and therefore, the sample for this project included the subsample from Congo. |
| EpiFloripa Aging Cohort Study | **EpiFloripa Idoso** | MMSE (≤ 17 ***) | The EpiFloripa Aging Cohort Study was carried out in the urban area of Florianópolis, State of Santa Catarina, located in the southern region of Brazil. The sample selection process was carried out by conglomerates in two steps. The units of the first step were the census tracts (IBGE census units) and the units of the second step were the households. Households were randomly selected. Mortality data linkage and active search for participants were used as follow-up strategies. |
| Etude Santé Psychologique et Traitement | **ESPRIT** | Standardized interview by a neurologist incorporating cognitive testing, with diagnoses validated by an independent panel of expert neurologists | The ESPRIT Study is a neuropsychiatric cohort study of community dwelling people aged 65 years and over drawn at random from the electoral rolls of Montpellier in the south of France and recruited between 1999 and 2001. |
| Invecchiamento Cerebrale in Abbiategrasso | **Invece.Ab** | DSM-IV | Eligible population comprises all people born between 1935 and 1939 who were residents living in Abbiategrasso, Milan, Italy on the start date of the study. All 1773 people born between 1935–39 residing in Abbiategrasso were contacted, a list obtained from the municipal registry office. Personalization of successive contacts were used. |
| Neuroprotective Model for Healthy | **LRGS TUA** | MMSE (≤14) | Population-based sample of Malaysian elderly from four states in Malaysia that have the highest numbers of older adults aged 60 years |

| | | | |
|---|---|---|---|
| Longevity among Malaysian Older Adults | | | and above, namely Johor, Perak, Selangor and Kelantan. The baseline study was conducted from May 2012 till February 2013, followed by 18 and 36 months follow-up. |
| Monongahela-Youghiogheny Healthy Aging Team | **MYHAT** | CDR (≥ 1) | An age-stratified sample of 2036 individuals aged 65+ years was drawn from the electoral rolls of a U.S. community, excluding individuals with severe cognitive impairment. |
| Sydney Memory and Ageing Study | **Sydney MAS** | DSM-IV | Non-demented community-dwelling individuals aged 70–90 were recruited from two areas of Sydney, following a random approach to 8914 individuals on the electoral roll. |
| Taiwan Initiative for Geriatric Epidemiological Research | **TIGER** | A history of dementia (self-report), medication use or MOCA-T (≤ 19) **** | Adults aged ≥ 65 years who participated in the senior health checkup program at National Taiwan University Hospital during 2011–2013 were recruited. |

*Participants with dementia at baseline were excluded from the analyses.

**For further details about each study, refer to the reference paper listed in Table 1.

***Dementia was not diagnosed in the study. The lowest 5th percentile (equivalent to a cut-off point of 17) is used to determine dementia, as recommended for the use in Brazilian population-based studies of elderly with low schooling level [Castro-Costa 2008].

**** MoCA-T = Montreal Cognitive Assessment-Taiwanese version. Dementia was not diagnosed in this study. There is no established cut-off for dementia for the MoCA-T and we excluded those in the lowest 5th percentile (equivalent to a cut-off point of 19) which is a conservative estimate for suspected dementia, as a large survey in Taiwan reported age-gender-adjusted prevalence of 8.1% for all cause dementia in adults aged 65+ [Sun 2014].

**eTable 2. Stroke data from each study**

| Study | Definition of incident stroke | Time of stroke (since study entry) |
|---|---|---|
| **EAS** | Self-reported, occurred in the past year | Time in study for the wave (which recorded the stroke) minus 0.5 |
| **EPIDEMCA** | Self-reported: Have you ever had an attack which requested medical attention? Or requiring hospitalization or consult due to stroke in the last 12 months | Time in study for the wave (which recorded the stroke) minus 0.5 |
| **EpiFloripa Idoso** | Self-reported: Has any doctor or health professional ever said that you have stroke | Halfway between two waves |
| **ESPRIT** | Self-reported | Time in study for the wave (which recorded the stroke) minus 1 year (for wave 4, minus 1.5) |
| **Invece.Ab** | World Health Organization (WHO) definition of stroke or TIA used | Year and month of stroke |
| **LRGS TUA** | Self-reported | Time since stroke (self-reported). If unavailable, half-way between two waves. |
| **MYHAT** | Self-reported | Halfway between two waves |
| **Sydney MAS** | Self-reported diagnosis of a stroke in the last 2 years (for wave 5: in the past 12 months) | >5 years ago, 1-2 years ago, 6-12 months ago, or 3-6 months ago |
| **TIGER** | Incident stroke self-reported at wave 3 and 4 (information was not collected at wave 2) | Halfway between two waves |

**eTable 3. Harmonization and definition of vascular risk factors from baseline**

| Study | Diabetes | Hypertension | High cholesterol | Cardiovascular disease | Depressive symptoms or depression |
|---|---|---|---|---|---|
| ***COSMIC harmonized criteria/ definition*** | **Medical history, treatment, self-report, and/or fasting blood glucose ≥126mg/dL or >7mmol/L** | **Medical history, treatment, self-report, and/or seated systolic blood pressure ≥140 mmHg or diastolic blood pressure ≥90 mmHg** | **Medical history, treatment, self-report, and/or total cholesterol ≥240mg/dL, or triglycerides ≥200mg/dL, or LDL ≥160 mg/dL** | **Atrial fibrillation, heart disease, angina, myocardial infarction, heart failure, and/or ischemic heart disease self-reported or in medical history notes** | **Anti-depressants, self-reported depression, and/or depression based on a scale and associated cut-off point** |
| EAS | Medical history | Blood pressure (mean of 2), medical history | Cholesterol, triglycerides | Myocardial infarction, coronary artery bypass, angina, heart failure, angioplasty, or arrhythmia | GDS score 6+ |
| EPIDEMCA | Medical history, treatment, blood glucose | Blood pressure (mean of 2), medical history, treatment | Cholesterol (>5.3mmol/L) | NA | GMS-AGECAT rating of subcase or clinical case |
| EpiFloripa Idoso | Self-report, fasting blood glucose | Blood pressure (mean of 2), self-report | Cholesterol, triglycerides, LDL | Self-report | Depression observed during neuropsychiatric exam |
| ESPRIT | Treatment, fasting blood glucose | Blood pressure (mean of 2), medication | Treatment, cholesterol, triglycerides | Ischemic heart disease (angina, history of angioplasty, heart operation or myocardial infarction), heartbeat disorders (arrhythmia or auricular fibrillation) | GDS score 6+; self-reported depression |
| Invece.Ab | Treatment, medical history | Blood pressure, medication | Medical history, treatment | Myocardial infarction, heart failure, angina, arrhythmia, coronary artery bypass graft, atrial fibrillation | Anti-depressants; GDS score 6+; Diagnosis by physician/ psychologist (medication, GDS score and CES-D items) |
| LRGS TUA | Self-reported | Self-reported or medication or blood pressure | Self-reported or medication | Heart disease | GDS score 6+ |
| MYHAT | Self-reported | Medical history, medication, blood pressure (average from waves 1 and 2) | Self-reported | Myocardial infarction, coronary heart failure | Use of anti-depressants; CES-D (modified) score ≥3 |
| Sydney MAS | Fasting blood glucose, treatment, medical history | Blood pressure (mean of 2), self-report, medication | Medical history, treatment, cholesterol, triglycerides | Heart attack, angina, cardiomyopathy, valve disease, arrhythmia, atrial fibrillation | GDS score 6+; use of medication |

| | | | | | |
|---|---|---|---|---|---|
| TIGER | Self-report, medication, fasting blood glucose | Self-report, medication | Cholesterol, self-report (for hyperlipidemia), medication | Coronary heart disease, atrial fibrillation (self-report) | Self-report; Medication use; CES-D score ≥ 16 |

Note. NA=not available. All medical history data were collected at baseline (wave 1). CES-D, Centre for Epidemiological Studies depression scale. CIRS, Cumulative Illness Rating Scale. DSM-IV, Diagnostic and Statistical Manual of Mental Disorders (4th edition). GDS, Geriatric Depression Scale. GMS-AGECAT, Geriatric Mental State-Automated Geriatric Examination for Computer Assisted Taxonomy. ICD-10, International Classification of Diseases (10th revision) MINI, Mini International Neuropsychiatric Interview, NPI, Neuropsychiatric Inventory.

**eTable 3. Harmonization and definition of vascular risk factors (continued from previous table)**

| Study | Alcohol use | Physical activity |
|---|---|---|
| **COSMIC** | **Nil/minimal=0; At least 1 drink per week = 1; 2+ drinks per week = 2** | **Minimally active = 0; Moderate activity at least once per week = 1; Vigorous activity at least once per week = 2.** |
| EAS | Drinks per week calculated from use during past year for each of beer, wine, liquor | Light activities (walking, gardening, dancing, calisthenics, golf, bowling, horse riding) at least 1 day a week = 1, Medium (hiking, tennis, cycling, swimming) or heavy (jogging, aerobics, hand, or racquet ball) activities at least 1 day a week=2, 0 (or less) days for all categories=0 |
| EPIDEMCA | Number of alcohol unit in a 'normal' week | Current physical activity was estimated using a thresh- old of at least 150 minutes of walking or cycling in the past week (World Health Organization 1984). |
| EpiFloripa Idoso | According to the AUDIT Alcohol screening tool. Never = 0; Moderate = 1; High = 2 | The physical activity score was measured using the long version of the International Physical Activity Questionnaire, adapted, and validated for the older adults in Brazil. The recreation and transportation domains were used. 1=≥150 min/week, 0=<150 min/week.[*] |
| ESPRIT | Drinks per week calculated from consumption in grams/day using 10 g = 1 drink | Either of gardening or walking “a little” or “a lot” = 1 vs “very little” = 0, Sports “regularly” or “often” = 2, gardening or walking “very little” and sports “never” or “from time to time” = 0 |
| Invece.Ab | Whether an individual drinks alcohol habitually, regardless of the amount. In Italy it is part of the normal diet to drink wine with meals. Data coded 0 or 1 only. | Response options are “never”, “1x week”, “2x week”, “3+ week”. Any of walking > 30 mins, dancing, “others”, group exercise “1x week” or more = 1. Any of cycling, swimming, running-jogging, tennis, aerobics “1x week” or more = 2. None of these or only walking < 30 mins = 0. |
| LRGS TUA | “How often do you drink alcoholic beverages?”<br>Never or 1 every month or less = 0; 2 to 4 times per month = 1; 2 to 3 times/week or more = 2 | NA |
| MYHAT | Calculated based on questions "have you ever had alcohol?", "How often do you drink?" and "How many drinks do you have at a time?" | Classed as bivariate |
| Sydney MAS | Coded as monthly or less = 0, 2-4 times a month = 1, 2-3 times a week or more = 2 | Any time indicated for separate activities bowls, golf, dancing, walking, other (Pilates, yoga, tai chi, weights)=1. Any time indicated for tennis, swimming, jogging, bicycling, aerobics, other relevant=2. Cases with no time indicated or where the time is clearly less than once a week=0. |
| TIGER | Never = 0; Ever = 1 | A short version of International Physical Activity Questionnaire (IPAQ) [**] was used to measure physical activity in MET minutes per week. ≥1500 MET-min/week = 2; ≥600 and <1500 MET-min/week = 1; <600 MET-min/week = 0. |

Note. NA=not available. [*] Ono LM et al 2015. [**] International Physical Activity Questionnaire.

**eTable 4. Neuropsychological tests used in each study for each cognitive domain**

| Study | Memory | Language | Processing Speed | Executive Function | Mini-Mental State Examination (MMSE) | Global Cognition* |
|---|---|---|---|---|---|---|
| **EAS** | Free and Cued Selective Reminding Test | Animals in 60s | TMTA | TMTB | Converted from Blessed | Yes |
| **EPIDEMCA** | Free and Cued Selective Reminding Test | Animals/ fruits in 60s | Zazzo's Cancellation Task | *NA* | *NA* | Yes |
| **EpiFloripa Idoso** | *NA* | *NA* | *NA* | *NA* | Yes | *NA* |
| **ESPRIT** | *Excluded* due to the *5-word test* having restricted 0–5 range and marked ceiling effects | Animals in 30s | TMTA | TMTB | Yes | Yes |
| **Invece.Ab** | Rey Auditory Verbal Learning Test, trial 7 (delay=15 min) | Verbal fluency, mean of colors/ animals/ fruits/ cities, each 120s | TMTA | TMTB | Yes | Yes |
| **LRGS TUA** | Rey Auditory Verbal Learning Test, trial 6 (delayed recall) | *NA* | *NA* | *NA* | Yes | *NA* |
| **MYHAT** | FULD | Animals in 60s | TMTA | TMTB | Yes | Yes |
| **Sydney MAS** | RAVLT | Animals in 60s | TMTA | TMTB | Yes | Yes |
| **TIGER** | Delayed free recall | Verbal fluency, mean of (fruits/fish/ vegetables) | TMTA | TMTB | NA | Yes |

Note. NA=not available. TMTA=Trail Making Test A; TMTB=Trail Making Test B. *Global cognition was calculated as the average of 3 or 4 domains.

**eTable 5. Participant baseline characteristics by study**

| Study | N | Age | Male | Education (years) | Mean FBG |
|---|---|---|---|---|---|
| **EAS** | 597 | 77.8 (5.3) | 219 (37) | 14.4 (3.4) | 5.52 (1.5) |
| **EPIDEMCA** | 184 | 74.8 (7.2) | 72 (39) | 2.5 (4.3) | 6.56 (1.9) |
| **EpiFloripa Idoso** | 542 | 68.8 (6.2) | 184 (34) | 8.3 (5.7) | 5.75 (1.7) |
| **ESPRIT** | 2060 | 73.0 (5.5) | 845 (41) | 10.2 (3.8) | 5.08 (1.1) |
| **Invece.Ab** | 1073 | 72.1 (6.4) | 490 (46) | 7.1 (3.3) | 6.21 (1.7) |
| **LRGS TUA** | 1702 | 68.5 (5.8) | 842 (49) | 5.3 (4.0) | 6.20 (2.2) |
| **MYHAT** | 458 | 74.6 (6.4) at baseline 81.6 (6.4) at wave which FBG was taken | 151 (33) | 13.2 (2.5) | 5.76 (1.7) |
| **Sydney MAS** | 896 | 78.6 (4.7) | 404 (45) | 11.7 (3.5) | 5.88 (1.2) |
| **TIGER** | 545 | 72.9 (5.4) | 256 (47) | 13.6 (3.7) | 5.56 (1.0) |
| **TOTAL** | 8,057 | 73.1 (6.5) Range=60 – 101 | 3463 (43) | 9.3 (5.0) | 5.74 (1.7) |

Note. Figures represent mean (SD) or n (%). FBG = fasting blood glucose level.

#### eTable 6. Missing data on covariates

| | Complete data for the full dataset | Imputed data for the full dataset |
|---|---|---|
| **Included in the adjusted model** | | |
| Smoking ever | 8051 | 6 |
| High cholesterol | 7969 | 88 |
| Depression | 7895 | 162 |
| Alcohol use | 6235 | 1822 |
| Physical activity | 6108 | 1949 |
| APOE4 carrier | 5469 | 2588 |
| **Not included in the adjusted model** | | |
| Hypertension | 8057 | 0 |
| Systolic blood pressure | 7933 | 124 |
| Diastolic blood pressure | 7934 | 124 |
| BMI | 7923 | 134 |
| CVD | 7869 | 188 |
| **For the primary analysis with global cognition data** | **Complete data for the primary analysis** | **Imputed data for the primary analysis** |
| Smoking ever | 5673 | 6 |
| High cholesterol | 5596 | 83 |
| Depression | 5557 | 122 |
| Alcohol use | 5490 | 189 |
| Physical activity | 5435 | 244 |
| APOE4 carrier | 5347 | 332 |

#### eTable 7. Mean values of covariates included in the adjusted model

| Covariates | Mean value* |
|---|---|
| Age at baseline (years) | 74.8 |
| Male sex (%) | 42% |
| Education (years) | 10.7 |
| White participants | 84% |
| High cholesterol (%) | 49% |
| APOE4 allele carrier (%) | 20% |
| Depressive symptoms (%) | 28% |
| Smoking, ever (%) | 42% |
| Physical activity – moderate and vigorous activity at least once a week (%) | 79% |
| Alcohol use – 1 or more drinks per week (%) | 59% |

Note. *Mean values of the subsample with global cognition data at baseline.

**eTable 8. Ethics approval of contributing cohorts**

| Study | Institutional Review Board |
|---|---|
| **EAS** | Albert Einstein College of Medicine Institutional Review Board (Approval#1996-175) |
| **EPIDEMCA** | Approved by Congolese ethical committee CERSSA (Comité d'Ethique de la Recherche en Sciences de Santé) and by an ethics review board (Comité de Protection des Personnes Sud-Ouest Outre Mer) in France. |
| **EpiFloripa Idoso** | Research Ethics Committee with Humans (CEPSH) of the Universidade Federal de Santa Catarina (UFSC), protocol 352/2008. In 2013/2014, it was approved by the Ethics Committee under CAAE 16731313.0.0000.0121. |
| **ESPRIT** | Ethics committee (CCPPRB) of the Kremlin Bicetre hospital (n° registered 99-28) |
| **Invece.Ab** | Ethics Committee of the University of Pavia (#3/2009) |
| **LRGS TUA** | Research and Medical Research Ethics Committee of Universiti Kebangsaan Malaysia (UKM) |
| **MYHAT** | Approved by the University of Pittsburgh Institutional Review Board |
| **Sydney MAS** | University of New South Wales Human Research Ethics Committee (approval #14327) |
| **TIGER** | Approved by the NTUH Research Ethics Committee (201101039RB, 201112047RIB, 201412213RINC, IRB 201712220RIN, 202012214RIN, 202112042RINA; 201312156RINC, 201712218RIN, 201812102RIN, 202012285RIN, 202112042RINA, and 202312052RINC) |

**eTable 9. Baseline characteristics of participants followed up until the last assessment* versus participants who dropped out**

| | Completed follow-up | Dropped out | P-value | Cohen's d/ Cohen's h |
|---|---|---|---|---|
| N | 4059 | 3998 | | |
| Age, y | 71.7 (6.0) | 73.5 (6.4) | <0.001 | 0.29 |
| Male sex, % | 1657 (41) | 1806 (44) | 0.006 | 0.06 |
| Education, y | 9.8 (4.8) | 8.9 (5.0) | <0.001 | 0.20 |
| White participants | 2366 (59) | 2483 (61) | 0.068 | 0.04 |
| BMI | 26.4 (4.6) | 25.7 (4.7) | <0.001 | 0.15 |
| APOE4 carrier | 535 (19) | 564 (21) | 0.20 | 0.05 |
| Blood pressure | | | | |
| Systolic | 138 (18) | 141 (20) | <0.001 | 0.15 |
| Diastolic | 78.6 (11) | 79.1 (11) | 0.028 | 0.05 |
| Hypertension | 2778 (69) | 2968 (73) | <0.001 | 0.09 |
| High cholesterol | 1842 (46) | 1602 (40) | <0.001 | 0.12 |
| Cardiovascular disease | 757 (19) | 875 (21) | <0.001 | 0.10 |
| Smoker (ever) | 1503 (38) | 1656 (41) | 0.003 | 0.05 |
| Alcohol use (at least one drink p/w) | 1716 (53) | 1820 (60) | <0.001 | 0.14 |
| Physical activity (moderate/vigorous) | 2469 (78) | 2166 (73) | <0.001 | 0.11 |
| History of depression | 785 (20) | 984 (25) | <0.001 | 0.12 |
| Baseline cognitive scores, SD | | | | |
| Global | 0.18 (1.0) | -0.12 (1.0) | <0.001 | 0.30 |
| Processing speed | 0.17 (1.0) | -0.11 (1.1) | <0.001 | 0.26 |
| Memory | 0.001 (1.1) | -0.28 (1.1) | <0.001 | 0.26 |
| Language | 0.16 (1.1) | -0.09 (1.1) | <0.001 | 0.22 |
| Executive function | 0.19 (1.10) | -0.10 (1.1) | <0.001 | 0.26 |
| Baseline MMSE (points) | 26.9 (3.1) | 26.0 (3.6) | <0.001 | 0.27 |

Note. Figures are mean (SD) or n (%). BMI= body mass index; and IFG= impaired fasting glucose.

Pearson $\chi 2$ tests, ANOVA, and t tests were used to examine group differences.

*Last assessment for the study or wave 7 for studies that followed up for greater than 11 years.

**eTable 10. Minimally adjusted and fully adjusted estimates of differences in baseline level and rate of decline in global cognition over time by glucose status group**

| | ***Basic model*** **(minimally adjusted)** | | ***Final fully adjusted model*** **(reproduced from Table 3 for comparison)** | |
|---|---|---|---|---|
| | **Coefficient (95% CI)** | **P-value** | **Coefficient (95% CI)** | **P-value** |
| ***Stroke-free follow-up*** | | | | |
| Baseline level<br>IFG vs Normal<br>T2D vs Normal | <br>-0.028 (-0.093, 0.038)<br>-0.16 (-0.23, -0.087) | <br>0.41<br>**<0.001** | <br>-0.029 (-0.094, 0.036)<br>-0.14 (-0.21, -0.069) | <br>0.38<br>**<0.001** |
| Slope (TIS) | -0.059 (-0.062, -0.056) | **<0.001** | -0.059 (-0.062, -0.056) | **<0.001** |
| TIS × IFG<br>TIS × T2D | 0.002 (-0.005, 0.008)<br>-0.004 (-0.011, 0.003) | 0.62<br>0.29 | 0.002 (-0.005, 0.008)<br>-0.004 (-0.012, 0.003) | 0.63<br>0.27 |
| ***Post-stroke*** | | | | |
| Acute effect of stroke on cognitive level (stroke) | -0.25 (-0.36, -0.15) | **<0.001** | -0.25 (-0.35, -0.15) | **<0.001** |
| Difference in slope relative to TIS (TSS) | -0.025 (-0.047, -0.002) | **0.036** | -0.025 (-0.047, -0.002) | **0.033** |

N=5631 (7 studies). T2D=Type 2 diabetes; IFG=impaired fasting glucose; normal=normal fasting glucose levels; TIS=time in study; TSS=time since stroke. Both models include covariates above plus age, sex, education, TIS × glucose group × age, and TIS × sex.

**eTable 11. Examination of baseline factors individually in the basic model**

| Characteristics (n) | Effect size (95% CI) | P-value |
|---|---|---|
| BMI (5512) | 0.0004 (-0.005, 0.006) | 0.89 |
| APOE ε4 carrier (5299) | -0.13 (-0.19, -0.07) | <0.001* |
| Blood pressure (5560) | | |
| Systolic | -0.0007 (-0.002, 0.001) | 0.34 |
| Diastolic | -0.0007 (-0.003, 0.002) | 0.58 |
| Hypertension (5631) | -0.018 (-0.070, 0.034) | 0.49 |
| High cholesterol (5548) | 0.051 (0.001, 0.10) | 0.044* |
| Cardiovascular disease (5466) | -0.022 (-0.079, 0.034) | 0.44 |
| Ever smoker (5625) | 0.060 (0.008, 0.11) | 0.023* |
| Alcohol use (5442) | | |
| Nil/minimal | Reference | |
| >=1 drink/week | 0.083 (0.029, 0.14) | 0.003* |
| Physical activity (5392) | | |
| Minimal | Reference | |
| Moderate/Vigorous | 0.083 (0.023, 0.14) | 0.007* |
| Depression (5511) | -0.22 (-0.28, -0.17) | <0.001* |

Note. n = participants with available data for global cognition. All characteristics represent measurements at study baseline or a medical history. *Significance at 0.1 level and the associated factor was chosen to be included in the fully adjusted model. The basic model included time in study (TIS), stroke, time since stroke (TSS), glucose status groups, age, sex, education, and interactions: group × TIS, age × TIS, and sex × TIS.

**eTable 12. Estimates of change in global cognition with interactions of post-stroke decline with glucose group**

| | **Coefficient (95% CI)** | **P-value** |
|---|---|---|
| ***Stroke-free follow-up*** | | |
| Baseline level<br>IFG vs Normal<br>T2D vs Normal | <br>-0.028 (-0.094, 0.038)<br>-0.16 (-0.23, -0.087) | <br>0.41<br>**<0.001** |
| Slope (TIS) | -0.065 (-0.067, -0.062) | **<0.001** |
| TIS × IFG<br>TIS × T2D | 0.0008 (-0.006, 0.007)<br>-0.0042 (-0.012, 0.003) | 0.82<br>0.28 |
| ***Post-stroke*** | | |
| Acute effect of stroke on cognitive level (stroke) | -0.32 (-0.46, -0.19) | **<0.001** |
| Difference in slope relative to TIS (TSS) | -0.019 (-0.046, 0.009) | 0.18 |
| Interactions<br>Stroke × IFG<br>Stroke × T2D<br>TSS × IFG<br>TSS × T2D | <br>0.24 (-0.021, 0.51)<br>0.078 (-0.17, 0.33)<br>-0.016 (-0.089, 0.050)<br>0.020 (-0.067, 0.053) | <br>0.072<br>0.54<br>0.58<br>0.82 |

Note. N=5631 (7 studies). T2D=Type 2 diabetes; IFG=impaired fasting glucose; normal=normal fasting glucose levels; TIS=time in study; TSS=time since stroke. The basic model included covariates above plus age, sex, education, TIS × age, TIS × sex.

**eTable 13. Examination of moderation effect of incident stroke status on cognitive decline over the stroke-free period**

| | Basic model with stroke status × TIS | | Basic model with stroke status × TIS × group | |
|---|---|---|---|---|
| **Measure** | **Coefficient (95% CI)** | **P-value** | **Coefficient (95% CI)** | **P-value** |
| Slope pre-/non-stroke (TIS; SD/y) | -0.063 (-0.066, -0.060) | <0.001 | -0.063 (-0.066, -0.060) | <0.001 |
| Baseline level | | | | |
| IFG | -0.026 (-0.095, 0.042) | 0.45 | -0.026 (-0.095, 0.044) | 0.47 |
| T2D | -0.15 (-0.22, -0.077) | <0.001 | -0.16 (-0.24, -0.086) | <0.001 |
| Baseline level | | | | |
| No stroke | Reference | - | Reference | - |
| Stroke | -0.046 (-0.17, 0.079) | 0.47 | -0.046 (-0.17, 0.079) | 0.47 |
| TIS × IFG | 0.003 (-0.004, 0.009) | 0.43 | 0.004 (-0.003, 0.010) | 0.31 |
| TIS × T2D | -0.003 (-0.010, 0.005) | 0.50 | -0.001 (-0.009, 0.007) | 0.76 |
| TIS × stroke status | -0.002 (-0.017, 0.013) | 0.80 | -0.002 (-0.017, 0.013) | 0.59 |
| Stroke status | Not included | | | |
| × IFG | | | -0.015 (-0.33, 0.30) | 0.93 |
| × T2D | | | 0.17 (-0.13, 0.48) | 0.27 |
| TIS × stroke status | Not included | | | |
| × IFG | | | -0.015 (-0.042, 0.012) | 0.28 |
| × T2D | | | -0.017 (-0.043, 0.009) | 0.20 |

Note. N=5631 (7 studies). Stroke = participant experienced first incident stroke over follow up. TIS=time in study; T2D=Type 2 diabetes; IFG=impaired fasting glucose; normal fasting glucose is the reference group. The models included covariates listed above plus age, sex, education, TIS × age, and TIS × sex, stroke (acute level change) and TSS (time after stroke).

**eTable 14. Sensitivity analyses**

| | **1: Excluded 2 studies with glucose measurements taken not at wave 1 N=5049 (5 studies)** | **2: Using complete data only N=4850 (7 studies)** |
|---|---|---|
| **Measure** | **Coefficient (95% CI); p-value** | **Coefficient (95% CI); p-value** |
| ***Stroke-free follow-up*** | | |
| Baseline level | | |
| IFG (vs Normal) | -0.038 (-0.10, 0.028); 0.26 | -0.018 (-0.087, 0.051); 0.61 |
| T2D (vs Normal) | -0.16 (-0.24, -0.090); **<0.001** | -0.13 (-0.21, -0.053); **0.001** |
| Slope (TIS) | -0.058 (-0.062, -0.055); **<0.001** | -0.060 (-0.063, -0.057); **<0.001** |
| TIS × IFG | 0.001 (-0.006, 0.008); 0.77 | 0.004 (-0.003, 0.011); 0.30 |
| TIS × T2D | -0.005 (-0.012, 0.003); 0.25 | -0.005 (-0.012, 0.003); 0.26 |
| ***Post-stroke*** | | |
| Acute effect of stroke (stroke) | -0.27 (-0.37, -0.16); **<0.001** | -0.32 (-0.43, -0.21); **<0.001** |
| Difference in slope relative to TIS (TSS) | -0.020 (-0.044, 0.003); 0.093 | -0.021 (-0.047, 0.004); 0.093 |

Note. T2D=Type 2 diabetes; IFG=impaired fasting glucose; normal=normal fasting glucose.
The fully adjusted models included the above covariates plus age, sex, education, race, high cholesterol, APOE4, history of depression, physical activity, alcohol use, smoking, and interactions age × TIS, sex × age.

#### eTable 15. Examination of cognitive domains and MMSE as outcome

| | **Coefficient (95% CI); p-value** | **Coefficient (95% CI); p-value** |
|---|---|---|
| | **Processing speed (SD)** | **Memory (SD)** |
| N (study) | 5700 (7) | 5280 (7) |
| Baseline level<br>IFG vs Normal<br>T2D vs Normal | <br>-0.031 (-0.10, 0.037); 0.37<br>-0.14 (-0.22, -0.069); **<0.001** | <br>-0.028 (-0.10, 0.045); 0.45<br>-0.10 (-0.17, -0.031); **0.005** |
| Slope (TIS) | -0.049 (-0.052, -0.044); **<0.001** | -0.036 (-0.043, -0.030); <0.001 |
| TIS × IFG<br>TIS × T2D | 0.0029 (-0.005, 0.011); 0.49<br>-0.0070 (-0.017, 0.0022); 0.13 | 0.006 (-0.005, 0.017); 0.30<br>-0.0004 (-0.012, 0.011); 0.95 |
| | **Language (SD)** | **Executive Function (SD)** |
| N (study) | 5719 (7) | 5346 (6) |
| Baseline level<br>IFG vs Normal<br>T2D vs Normal | <br>-0.015 (-0.085, 0.055); 0.67<br>-0.095 (-0.17, -0.020); **0.013** | <br>-0.004 (-0.077, 0.068); 0.91<br>-0.14 (-0.22, -0.061); **0.001** |
| Slope (TIS) | -0.044 (-0.047, -0.040); **<0.001** | -0.070 (-0.074, -0.066); **<0.001** |
| TIS × IFG<br>TIS × T2D | -0.0003 (-0.008, 0.008); 0.95<br>-0.002 (-0.011, 0.006); 0.61 | 0.0005 (-0.008, 0.009); 0.92<br>-0.004 (-0.014, 0.006); 0.43 |
| | **MMSE (raw score)** | |
| N (study) | 7321 (7) | |
| Baseline level<br>IFG vs Normal<br>T2D vs Normal | <br>0.080 (-0.085, 0.24); 0.35<br>-0.044 (-0.21, 0.12); 0.60 | |
| Slope (TIS) | -0.046 (-0.056, -0.036); **<0.001** | |
| TIS × IFG<br>TIS × T2D | -0.054 (-0.073, -0.034); **<0.001**<br>-0.053 (-0.075, -0.032); **<0.001** | |

Note. T2D=Type 2 diabetes; IFG=impaired fasting glucose; normal=normal fasting glucose. Normal fasting glucose is the reference group. The fully adjusted models included post-stroke effects TSS, stroke, and covariates baseline age, sex, education, race, high cholesterol, APOE4 carrier, history of depression, physical activity, alcohol use, smoking, and interactions age × TIS, sex × age.

##### eTable 16. Subgroup analysis 1 – cohorts with follow-up fasting glucose measurements

##### eTable 16A. Studies with follow-up fasting glucose measurement

| Study | Country | Cognitive assessment | Waves | N |
|---|---|---|---|---|
| EAS | USA | Global cognition | 1, 2 | 398 |
| EPIDEMCA | Republic of Congo | Global cognition | 2, 3 | 108 |
| Invece.Ab | Italy | Global cognition | 1, 2 | 917 |
| Sydney MAS | Australia | Global cognition | 1, 2 | 711 |

##### eTable 16B. Defining change in glucose status groups in studies with global cognition

| Change in glucose status groups | N (%) |
|---|---|
| 1 Stable normal (normal to normal) | 709 (33) |
| 2 Stable IFG (IFG to IFG) | 415 (19) |
| 3 IFG to normal | 206 (9.7) |
| 4 IFG to T2D | 33 (1.6) |
| 5 Incident glucose disorders (normal to IFG or T2D) | 197 (9.2) |
| 6 Baseline T2D (regardless of change) | 574 (27) |
| **Total** | 2134 |

Note. Normal = normal fasting glucose; IFG=impaired fasting glucose; T2D= type 2 diabetes.

##### eTable 16C. Estimates of cognitive decline in global cognition

| | **In all participants with available data N=2134 (4 studies)** | | **Excluding cognitive assessment prior to glucose collection * N =1995 (4 studies)** | |
|---|---|---|---|---|
| | **Coefficient (95% CI)** | **P-value** | **Coefficient (95% CI)** | **P-value** |
| ***Stroke-free follow-up*** | | | | |
| Baseline level | | | | |
| Stable normal | Ref | | Ref | |
| Stable IFG | -0.022 (-0.14, 0.09) | 0.71 | 0.006 (-0.13, 0.14) | 0.94 |
| IFG to normal | -0.032 (-0.18, 0.11) | 0.66 | -0.016 (-0.19, 0.16) | 0.86 |
| IFG to T2D | -0.076 (-0.40, 0.25) | 0.64 | -0.092 (-0.47, 0.29) | 0.63 |
| Normal to IFG or T2D | 0.061 (-0.09, 0.21) | 0.42 | 0.12 (-0.054, 0.29) | 0.18 |
| Baseline T2D | -0.19 (-0.29, -0.08) | **0.001** | -0.12 (-0.24, 0.011) | 0.074 |
| Slope (TIS; SD/y) | -0.070 (-0.078, -0.062) | **<0.001** | -0.070 (-0.081, -0.060) | **<0.001** |
| TIS × | | | | |
| Stable normal | Ref | | Ref | |
| Stable IFG | 0.007 (-0.006, 0.02) | 0.30 | 0.004 (-0.014, 0.022) | 0.65 |
| IFG to normal | -0.008 (-0.03, 0.01) | 0.36 | -0.009 (-0.034, 0.017) | 0.50 |
| IFG to T2D | 0.023 (-0.012, 0.06) | 0.20 | 0.021 (-0.028, 0.070) | 0.39 |
| Normal to IFG or T2D | -0.005 (-0.021, 0.011) | 0.54 | -0.011 (-0.033, 0.010) | 0.31 |
| Baseline T2D | 0.003 (-0.009, 0.016) | 0.62 | -0.002 (-0.020, 0.015) | 0.81 |

Note. T2D=Type 2 diabetes; IFG=impaired fasting glucose; normal=normal fasting glucose levels. The fully adjusted models included post-stroke effects TSS, stroke, and adjusted for baseline age, sex, education, race, high cholesterol, APOE4 carrier, history of depression, physical activity, alcohol use, smoking, and interactions age × TIS, sex × age.

**eTable 17. Subgroup analysis 2 – Average fasting glucose levels to define glucose groups in studies with follow-up glucose measurements**

| **Global cognition** | **In all participants with available data N =1963 (4 studies)** | | **Excluding cognitive assessment prior to glucose collection * N =1894 (4 studies)** | |
|---|---|---|---|---|
| ***Stroke-free follow-up*** | **Coefficient (95% CI)** | **P-value** | **Coefficient (95% CI)** | **P-value** |
| Baseline level | | | | |
| IFG | 0.016 (-0.080, 0.11) | 0.75 | 0.036 (-0.077, 0.15) | 0.53 |
| T2D | -0.12 (-0.23, -0.010) | **0.032** | -0.10 (-0.23, 0.022) | 0.11 |
| Slope (TIS) | -0.071 (-0.078, -0.064) | **<0.001** | -0.072 (-0.081, -0.062) | **<0.001** |
| TIS × IFG | 0.004 (-0.007, 0.015) | 0.45 | 0.002 (-0.013, 0.017) | 0.79 |
| TIS × T2D | 0.001 (-0.012, 0.014) | 0.88 | -0.002 (-0.020, 0.015) | 0.79 |

Note. T2D=Type 2 diabetes; IFG=impaired fasting glucose; normal fasting glucose levels is the reference group. The fully adjusted models included post-stroke effects TSS, stroke, age, sex, education, race, high cholesterol, APOE4 carrier, history of depression, physical activity, alcohol use, smoking, and interactions age × TIS, and sex × age.

#### eTable 18. Subgroup analysis 3 – cohorts with metformin data

#### eTable 18A. Studies with metformin data and global cognition

| Study | Country | N |
|---|---|---|
| EAS | USA | 591 |
| MYHAT | USA | 386 |
| Sydney MAS | Australia | 884 |
| TIGER | Taiwan | 545 |

Note. One study (EpiFloripa) collected fasting glucose measurement at follow-up but did not have global cognition available and was therefore not included in this analysis.

#### eTable 18B. Estimates of change in global cognition by glucose status and metformin groups

| Measure (model variable) | Coefficient (95% CI) | P-value |
|---|---|---|
| ***Stroke-free follow-up*** | | |
| Baseline level (group) | | |
| IFG | 0.063 (-0.09, 0.21) | 0.42 |
| T2D with metformin | -0.19 (-0.32, -0.059) | **0.004** |
| T2D without metformin | -0.14 (-0.27, -0.013) | **0.031** |
| Slope (TIS; SD/y) | -0.047 (-0.053, -0.041) | **<0.001** |
| TIS × IFG | 0.010 (-0.009, 0.030) | 0.29 |
| TIS × T2D with metformin | 0.010 (-0.006, 0.025) | 0.22 |
| TIS × T2D without metformin | 0.0011 (-0.016, 0.018) | 0.91 |

N=2441 from 4 studies. Group=glucose status group; TIS=time in study; TSS=time since stroke; T2D=type 2 diabetes; IFG=impaired fasting glucose; normal=normal fasting glucose levels (reference group). The fully adjusted model included age, sex, education, race, high cholesterol, APOE4, smoking, alcohol use, physical activity, history of depression, interactions age × TIS, and sex × age. Bold figures indicate p<0.05.

### SUPPLEMENTARY FIGURES

**eFigure 1. Follow-up schedule and numbers of participants for each contributing study cohort**

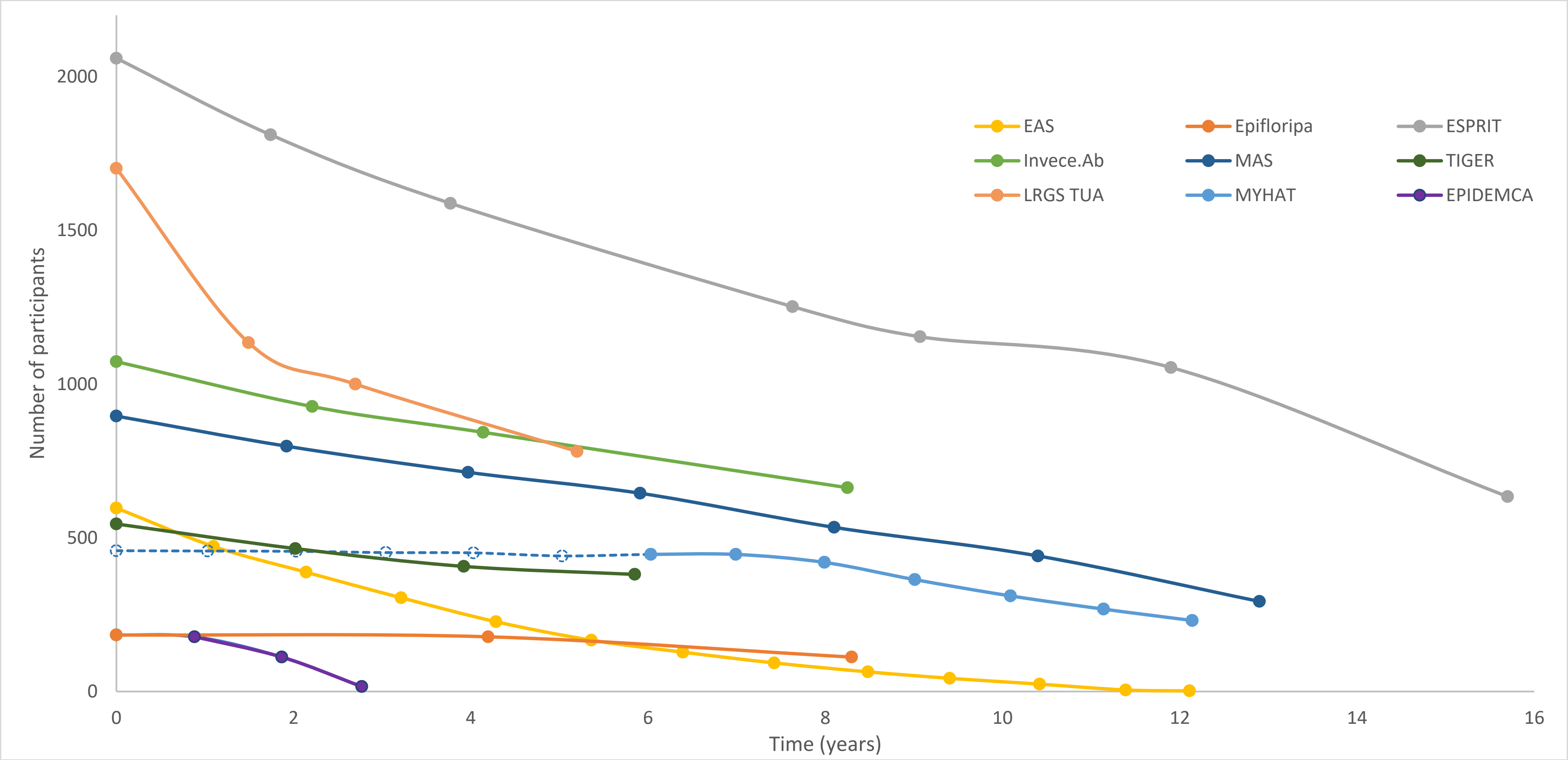

Note. Dotted line indicates cognitive assessments conducted prior to collection of fasting glucose measurements and were not included in the analyses.

**eFigure 2. Trajectory of cognitive decline in cognitive domain z-scores and MMSE by glucose status groups**

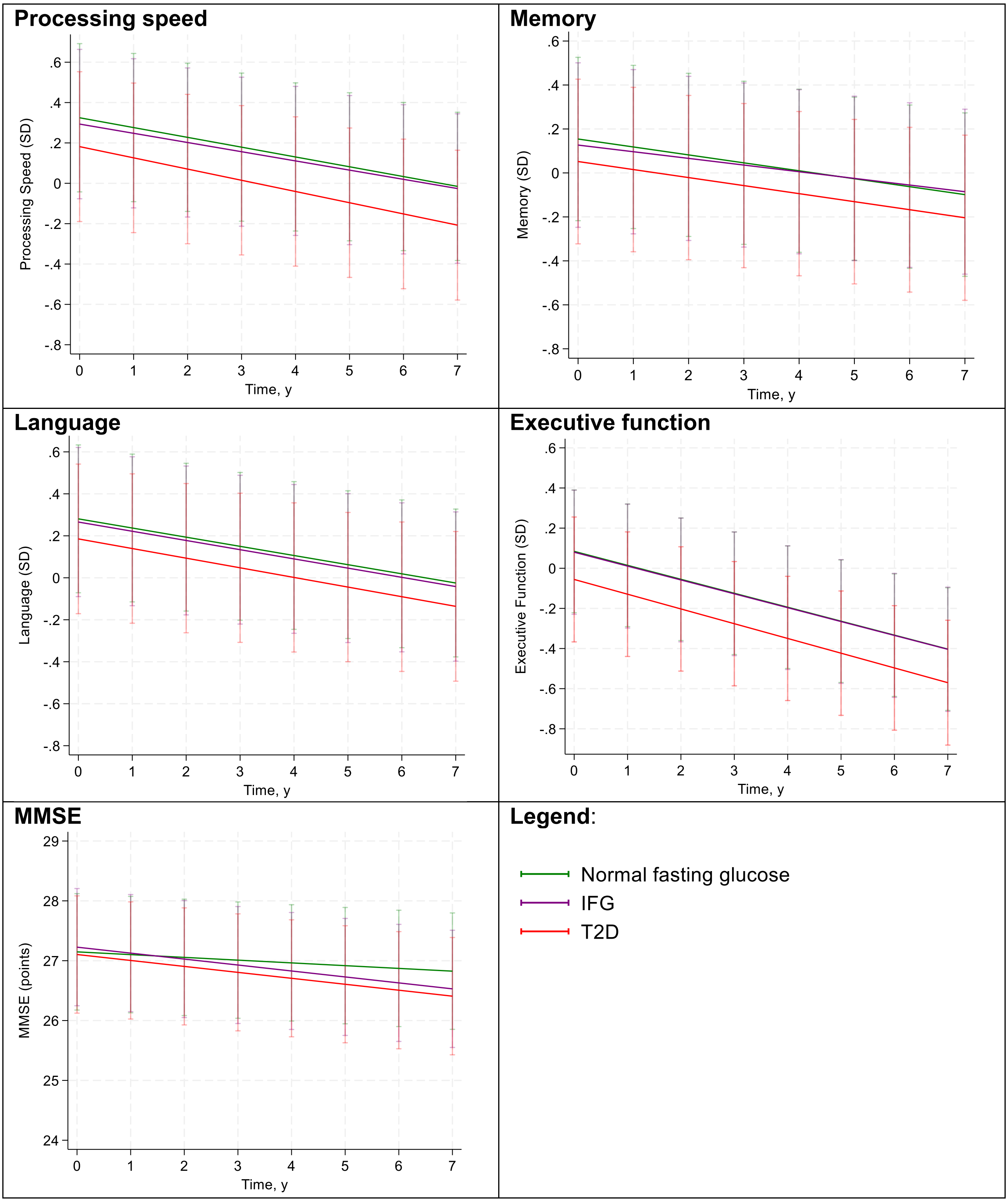

Note. Trajectories were plotted over the stroke-free period. Predicted values of cognition scores were calculated for common values of covariates at baseline. Common values were based on subsample with global cognition data, see eTable 7. Vertical bars indicate 95% CI.
